# Cost-Effectiveness of Continuous Glucose Monitoring for people with elevated HbA1c living with Type 1 Diabetes in South Africa: A Within-Trial and Modeled Economic Evaluation

**DOI:** 10.64898/2026.07.22.26358646

**Authors:** Sarah Girdwood, Elena Marbán-Castro, Lorrein Muhwava, Janetta Catharina de Beer, Cathy Haldane, Joel A Dave, Michelle Carrihill, Maria Karsas, Paul Rheeder

**Affiliations:** FIND, Geneva, Switzerland; VI Research, Centurion, South Africa; Division of Endocrinology, Groote Schuur Hospital and the University of Cape Town, Cape Town, South Africa; Red Cross Childrens Hospital, Paediatric Clinic, Cape Town, South Africa; Departments of Paediatrics and Internal Medicine, Steve Biko Academic Hospital, University of Pretoria, Pretoria, South Africa

## Abstract

**Background:** Continuous glucose monitoring (CGM) improves glycaemic control in people with type 1 diabetes (T1D), but high costs limit uptake in low- and middle-income (LMIC) countries. Evidence on the cost-effectiveness of CGM is limited in LMIC settings.

**Objective:** To evaluate the short- and long-term cost-effectiveness of intermittently-scanned continuous CGM (cCGM) and intermittently-scanned periodic CGM (pCGM) (one sensor every three months), versus standard self-monitoring of blood glucose (SMBG) in the South African public-sector.

**Methods:** A three-arm randomised controlled trial (ACCEDE) was conducted among T1D individuals with HbA1c ≥10% in South Africa. A within-trial cost-effectiveness analysis was conducted from a partial societal perspective over 9-months, using resource use data and QALYs derived from EQ-5D. A cost-utility analysis using a Markov microsimulation model was conducted for two populations: total T1D, and youth (<20 years).

**Results:** Within-trial analysis showed no statistically significant differences in QALYs or HbA1c between arms. Costs were highest for cCGM (USD 1,504), followed by pCGM (USD 742) and SMBG (USD 467). CGM strategies were dominated in the within-trial analysis. In contrast, long-term modelling showed that CGM was more effective than SMBG and was cost-effective for youth when used periodically. cCGM delivered additional QALYs at a higher cost (ICERs USD 15,259-30,852/QALY) and was only potentially cost-effective in youth when sensor prices were reduced by >45%.

**Conclusions:** While CGM was not cost-effective in the short term, modelling suggests pCGM use may offer value-for-money under specific assumptions and in select populations, highlighting the need for further evidence on long-term effectiveness, engagement, and pricing.

## Introduction

People living with Type 1 diabetes (T1D) require lifelong, consistent monitoring of blood glucose levels to prevent both acute and long-term complications. Self-monitoring of blood glucose (SMBG) has long been a cornerstone of diabetes management, enabling individuals to make informed adjustments to their diet, physical activity, and medication. SMBG typically involves capillary fingerstick testing multiple times a day; however, this approach is often painful and burdensome, leading to poor adherence and suboptimal glycaemic control. Moreover, SMBG provides only intermittent snapshots of glucose levels, which can result in missed or delayed detection of hypo- and hyperglycaemic episodes. Suboptimal glycaemic control significantly increases the risk of serious microvascular and macrovascular complications, which in turn contribute to the overall cost of diabetes care^1,2^.

In recent years, continuous glucose monitoring (CGM) technology has emerged as a transformative tool in diabetes care. CGMs offer real-time glucose readings, trend information, and alarms for hypo- or hyperglycaemia, providing patients and healthcare providers with far more comprehensive insights into glycemic patterns. Numerous studies have demonstrated that CGMs improve HbA1c, reduce the time spent in hypo- or hyperglycaemia, and enhance quality of life by minimizing reliance on fingerstick and reducing the fear of hypoglycaemia^3–11^. CGMs have shown particular benefit in people with insulin-treated type 1 and type 2 diabetes, both clinically and economically, with multiple evaluations supporting their cost-effectiveness^12–18^.

In high-income countries, CGMs are now widely accessible and reimbursed, resulting in widespread uptake among individuals with T1D^17^. However, in most low- and middle-income countries (LMICs), including South Africa, the standard of care for glucose monitoring still relies on finger-prick capillary testing, while access to CGM remains severely limited. High sensor costs, lack of reimbursement, and weak public-sector procurement mechanisms continue to hinder adoption^19^. In South Africa, while some progress has been made in the private sector, where CGMs are often partially or fully covered by insurance, individuals with T1D in the public sector rarely have access to this technology.

To address the evidence-gap in LMICs, we conducted a pragmatic randomised controlled trial (RCT) (ACCEDE, trial registry number NCT05944718) of intermittently-scanned continuous CGM (cCGM) and intermittently-scanned CGM used periodically (pCGM) in the public sector in South Africa, alongside a comprehensive economic evaluation. This paper presents the results of the within-trial and modeled cost-effectiveness analysis, estimating both short-term (trial-based) and long-term value for money of CGM in the South African health system.

## Methods

### Study design

A three-arm, pragmatic RCT was conducted at three public sector hospitals in South Africa (two based in Cape Town, and one in Pretoria). 248 participants were enrolled and included children (> 4 years), adolescents, and adults with T1D and baseline HbA1c ≥10%. Participants were randomised 1:1:1 to one of the following arms:

- ARM 1 - *cCGM* using Freestyle Libre sensors replaced every 14 days for 9 months;
- ARM 2 - *pCGM* using Freestyle Libre sensors for 2-week intervals every 3 months alternating with SMBG for 9 months;
- ARM 3 - *Standard Care*, consisting of SMBG for 9 months.

The intervention period lasted 9 months, during which participants in the intervention arms received CGMs and were followed up through structured study visits. While data were collected up to 15 months post-enrolment, there was no CGM intervention beyond 9 months. All participants received diabetes education and were surveyed at baseline, 3, 6, and 9 months regarding their health-related quality of life, healthcare service utilisation and work productivity losses if employed (absenteeism and presenteeism), and caregiver time devoted to caring for children with T1D. More details on the ACCEDE trial, including its design, study population and clinical results, can be found in Marbán-Castron et al, 2024^20^.

### Within-trial cost-effectiveness analysis

Our cost-effectiveness analysis (CEA) included a within-trial CEA using observed trial data conducted from a partial societal perspective, capturing both direct healthcare costs and indirect participant costs. The time horizon aligned with the 9-month duration of the trial.

#### Measurement of effectiveness

The primary effectiveness outcome was quality-adjusted life years (QALYs), estimated using baseline and follow-up utility scores derived from EQ-5D instruments. The EQ-5D-3L was used for adults (≥16 years), EQ-5D-Y for youth (8–15 years), and EQ-5D-Proxy for caregivers of younger children^21^. In the absence of a South African-specific value set, utility scores were derived using the Zimbabwean value set for adult and proxy data, and the Indonesian value set for youth reflecting the closest available geographical and socio-economic comparators^22,23^. As a sensitivity analysis, UK (adult/proxy) and Slovenian (youth) value sets were applied^24^. QALYs were calculated using the area under the curve over the trial period (median 9 months)^25^. QALYs and utility change were calculated from observed participant-level data. Between-arm differences were estimated using linear regression models adjusting for baseline utility, age, and EQ-5D instrument type, in line with recommended methods for trial-based economic evaluation^26,27^. Utility at death was set to 0 (n=3). Secondary outcomes included change in HbA1c from baseline. Descriptive statistics are presented as observed values, with between-arm comparisons based on adjusted model estimates.

#### Resource use and costs

Standard micro-costing methods were used to estimate the costs of both direct and indirect resources utilised by participants over the 9-month duration of the trial. Direct resource use was recorded retrospectively and included CGM usage (sensors, adhesive plasters and readers), glucose monitoring supplies (strips, lancets, glucometer), other diabetes-related consumables (needles, glucagon kits, syringes), medicines (including insulin), laboratory and other diagnostic tests, and healthcare utilisation outside of the trial (outpatient visits, inpatient admissions). Nurse educator visits were delivered to all participants irrespective of study arm and were included in the healthcare provider cost category. Trial-related procedures or research activities were excluded from the cost analysis.

Unit costs were assigned to these resource outputs using prices sourced from public sources, and where necessary the literature^28–33^. Unit costs for the Abbott Freestyle Libre 1 and 2 devices were obtained from trial records and reflect the full in-country price, inclusive of import duties and local distribution costs: USD 60.50 per sensor and USD 58.89 per reader. Additional detail on costing sources and assumptions is provided in Table S8 in the Supplementary Appendix.

The average cost per participant per Arm was determined by multiplying the resource usage with the associated unit costs for the 9-month duration of the trial. All values were reported in South African Rands (ZAR) and converted to United States Dollars (USD) at the rate of R18.00/USD (the average exchange rate over the period 1 January to 19 November 2025)^34^.

#### Cost-effectiveness analysis

The cost-effectiveness analysis was conducted as a within-trial analysis using the intention-to-treat results and presented in terms of incremental cost-effectiveness ratios (ICERs) which compare the additional costs and outcomes of each of the Arms relative to the standard of care. ICERs were calculated as the arithmetic mean difference in costs and QALYs between Arms.

### Long-term modelled analysis

#### Model overview

To complement the within-trial CEA, we conducted a lifetime cost-utility analysis using the open-source DEDUCE model (Determination of Diabetes Utilities, Costs, and Effects)^35^. The model is a Monte Carlo, individual patient-level microsimulation model specifically developed to evaluate diabetes interventions and incorporates glycemic trajectories, complication risks, and healthcare costs over a patient’s lifetime.

Building on observed trial data, including HbA1c reduction and baseline complication prevalence, the model simulates the long-term incidence of major diabetes-related complications (e.g., diabetic ketoacidosis (DKA), nephropathy, cardiovascular disease, retinopathy/blindness, and severe hypoglycaemia events (SHE)), associated healthcare costs, and quality-adjusted survival. Alongside HbA1c, acute event rates, specifically DKA and SHE hospitalisations, are key short-term drivers of model outcomes. Annual probabilities for DKA and SHE were derived directly from trial data by arm and sub-population and are reported in full in Supplementary Table 4. Transition probabilities were derived from published international risk equations - the Sheffield type 1 Diabetes Model equations ^36,37,38^. The model was localised with epidemiological and clinical patterns observed in the South African public sector context, where data was available.

The model uses annual cycles and allows for concurrent complications and mortality. Baseline health utilities were assigned based on the within-trial analysis (Supplementary Appendix Table S6). Utility decrements for complications were drawn from international literature and applied additively in the presence of multiple events^39,40,41,42,43,44^ (Supplementary Appendix Table S7). QALYs were calculated by applying health-state utility weights to simulated survival over each model cycle and aggregating these over the model time horizon.

#### Modelled populations and time horizon

To evaluate the long-term cost-effectiveness of CGM in different subpopulations, two cohort-specific instances of the DEDUCE model were constructed to assess whether CGM offers differential value for money in specific sub-populations. In each, either cCGM or pCGM use was compared against the standard of care, SMBG. The modeled cohorts included:

1. **Total population:** comprising all trial participants (adults and youth), weighted to reflect the age distribution observed in the trial.
2. **Youth**: individuals under the age of 20, including children and adolescents.

For the total population, the model horizon spanned a lifetime (100 years), consistent with similar long-term diabetes evaluations. As the DEDUCE model does not support age-related transitions in clinical parameters, the time horizon modelled for the Youth sub-population was 15. This horizon ensures the youngest participants (∼4 years at baseline) reach adulthood within the model period. This likely underestimates the long-term QALY gains from improved glycaemic control which are known to compound over decades.

#### Resource use and costs

Costs were estimated from a partial societal perspective, consistent with the within-trial CEA. Blood glucose monitoring testing frequency was determined by age group and trial arm (higher daily testing in children, reduced fingerstick use when participants were on CGM). CGM costs reflected sensor use as per trial protocols (continuous replacement every 14 days in Arm 1; 2 weeks of use every 3 months in Arm 2) along with one reader device per participant and the use of adhesive plasters for each sensor. As the DEDUCE model does not capture age-related transitions in productivity or caregiving, indirect costs were estimated using an event-based approach (Supplementary Appendix). Input prices were based on the same local sources as the trial and inflated to 2025 ZAR. Other longer-term costs that were not measured in the trial (complication costs) were taken from Bhana et al. 2023 or micro-costed (Supplementary Appendix Table S9)^45^.

#### Model assumptions

Future costs and QALYs were discounted at 5% annually^46^. Results are reported as ICERs in USD per QALY gained using the same conversion as the within-trial analysis (ZAR 18/USD). As South Africa does not have an explicitly adopted cost-effectiveness threshold for decision-making, results were interpreted against published estimates of opportunity cost–based thresholds for the South African health system. We used the upper-bound estimate proposed by Pichon-Rivière et al., corresponding to approximately USD 4,600 per QALY in 2025 (68% of per capita GDP), as the primary reference threshold^47^. For contextual comparison, we also present results across a broader range of willingness-to-pay (WTP) thresholds, including 1–3 times South Africa’s gross domestic product (GDP) per capita, consistent with historical WHO-CHOICE benchmarks. The model structure and analytic approach align with the CHEERS 2022 guidelines^48^ (Supplementary Appendix Table 11).

#### Sensitivity analysis

We conducted deterministic one-way sensitivity analyses to examine the impact of parameter uncertainty on the incremental cost-effectiveness results by varying structural, costing, and treatment effect parameters; a threshold analysis estimated the CGM price reduction required for cost-effectiveness at relevant WTP thresholds. Probabilistic sensitivity analysis (PSA) was undertaken using Monte Carlo simulation, drawing parameters from prespecified distributions reflecting parameter uncertainty (see Supplementary Appendix).

### Ethics

The protocol was approved by the Faculty of Health Sciences Research Ethics Committee at the University of Pretoria (330/2023) and the Human Research Ethics Committee at the University of Cape Town (HREC REF 558/2023). Written, informed consent to participate in the trial was obtained from all participants and/or their parents/caregivers.

### Patient involvement

Patients and the public were not involved in designing this economic evaluation. A lived experience group convened through SA Diabetes Advocacy advised on the design of the underlying ACCEDE trial and on how findings, including those reported here, are communicated.

## Results

### Baseline characteristics

A total of 248 participants were enrolled in the study, 245 of whom attended the first study visit, and a further 227 completed the 9-month end of intervention visit. Baseline characteristics were generally well balanced across the study arms, with no significant differences observed between intervention and control groups (Table 1). The only exception was the frequency of self-reported self-monitored fingerstick glucose testing, which was significantly higher in the arm assigned to pCGM (Arm 2).

**Table 1:** Baseline characteristics by arm.

| Variable<sup>1</sup> | N | ARM 1<br>N = 81 | ARM 2<br>N = 85 | ARM 3<br>N = 79 | p-value<sup>2</sup> |
| --- | --- | --- | --- | --- | --- |
| Age (years) | 245 | 20.46 (10.39) | 21.02 (12.07) | 22.46 (12.58) | 0.5 |
| Years since diagnosis | 245 | 9.38 (7.25) | 9.49 (7.46) | 10.75 (7.35) | 0.4 |
| Baseline HbA1c (%) | 240 | 12.39 (2.08) | 12.02 (1.59) | 12.16 (1.78) | 0.4 |
| Baseline SMBG frequency/day | 243 | 3.29 (1.19) | 3.69 (1.85) | 2.99 (1.30) | 0.010 |
| Female | 245 | 49 (60%) | 55 (65%) | 44 (56%) | 0.5 |

<sup>1</sup>Mean (SD); n / N (%)
<sup>2</sup>One-way analysis of means; Pearson's Chi-squared test

### Within trial CEA

QALYs and utility values were similar across trial arms, with no statistically significant differences in adjusted analyses (Arm 1 vs Arm 3: Δ −0.005; 95% CI −0.023 to 0.013; Arm 2 vs Arm 3: Δ 0.002; 95% CI −0.020 to 0.024). Results were consistent across alternative EQ-5D value sets (Supplementary Table S10). Full utility and QALY data are presented in Table 2.

**Table 2:**
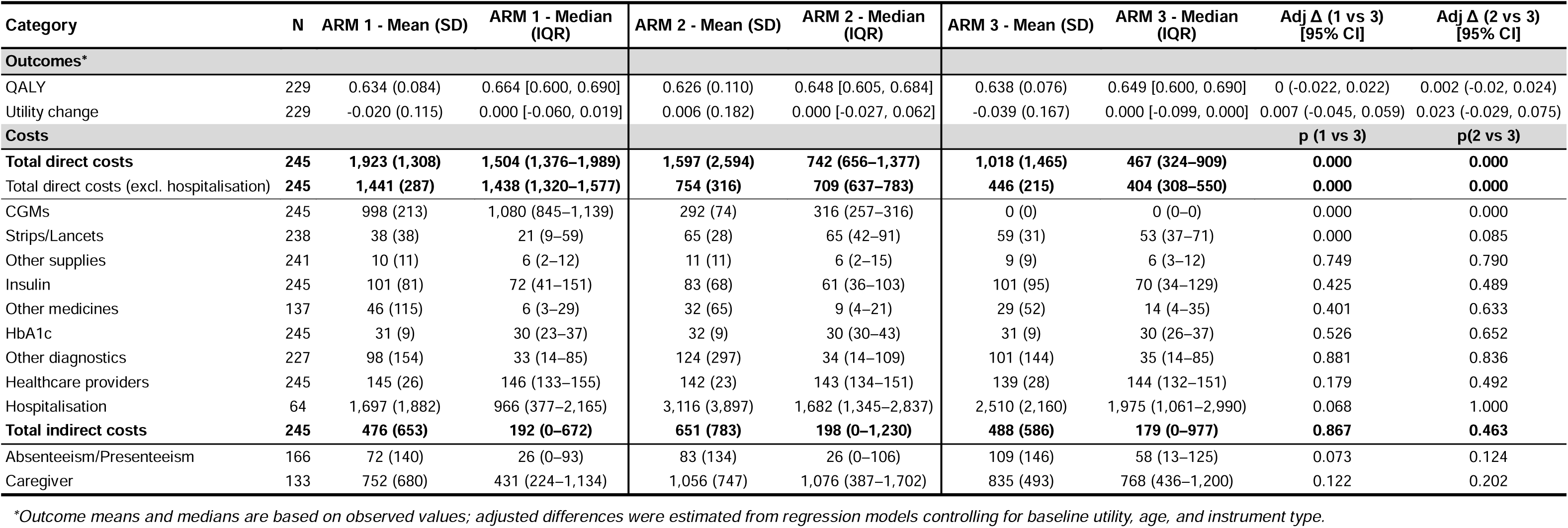
Within-trial results: Outcomes and costs by ARM: average cost per participant per ARM for the trial duration (USD)

From a partial societal perspective, the median 9-month total direct costs were highest in the cCGM arm (ARM 1; USD 1,504) compared with the pCGM arm (ARM 2; USD 742) and the control arm (ARM 3; USD 467; *p*<0.001) (Table 2). These differences were primarily attributable to CGM device expenditures (ARM 1: USD 1,080; ARM 2: USD 316; ARM 3: USD 0), with non-significant trends toward lower hospitalisation and absenteeism costs in the CGM arms; full cost breakdowns are presented in Table 2.

No within-trial ICER was calculated due to the lack of difference in QALYs. Both the cCGM (ARM 1) and the pCGM arm (ARM 2) were dominated by control in the within-trial CEA.

HbA1c, a key input for the long-term model, showed no statistically significant between-arm differences at month 9; exploratory subgroup analyses in youth and fully compliant participants were similarly non-significant (Supplementary Table S1). A larger and statistically significant HbA1c reduction was observed at month 6 for pCGM (Supplementary Table S1).

DKA was the leading cause of acute hospitalisation across all trial arms, with the annual probability in the SMBG arm reaching 31% in the total population and 46% in youth (Supplementary Table S4). CGM was associated with relative reductions of 34% (cCGM) and 9% (pCGM) in the total population, and 46% and 22% respectively in youth. SHE rates followed a similar pattern, with relative reductions of 67% and 22% for cCGM and pCGM in the total population, and 69% and 39% in youth.

### Long-term CEA

In the long-term model, CGM arms were more effective in terms of QALYs gained than SMBG (Arm 3), while cCGM (Arm 1) generated the largest QALY gains but at substantially higher incremental cost (Table 3). In the overall population, discounted lifetime costs were USD 34,626 for SMBG, USD 50,102 for cCGM, and USD 36,765 for pCGM. Compared with SMBG, cCGM yielded an additional 0.502 QALYs at an incremental cost of USD 15,476 (ICER: USD 30,852 per QALY gained). pCGM generated 0.038 additional QALYs at an incremental cost of USD 2,139, corresponding to an ICER of USD 56,128 per QALY gained.

**Table 3:** Long-term modelled cost-effectiveness of CGM strategies versus SMBG: all T1D individuals and youth subgroup.

| Cohort | Strategy | Total Costs (USD) | Life years gained | QALYs | Incremental Costs vs SMBG (USD) | Incremental QALYs vs SMBG | ICER (USD/ QALY) |
| --- | --- | --- | --- | --- | --- | --- | --- |
| All : lifetime horizon | Arm 3: SMBG | \$34 626 | 10.306 | 7.945 | | | |
| | Arm 1: CGM | \$50 102 | 10.937 | 8.447 | \$15 476 | 0.502 | \$30 852 |
| | Arm 2: CGM (Periodic) | \$36 765 | 10.382 | 7.983 | \$2 139 | 0.038 | \$56 128 |
| Youth (age < 20): 15 year time horizon | Arm 3: SMBG | \$27 362 | 8.098 | 6.584 | | | |

Results were more favourable for youth (<20 years), with pCGM producing 0.226 additional QALYs at an incremental cost of USD 540 relative to SMBG (ICER: USD 2,390 per QALY gained). cCGM in youth yielded 0.604 additional QALYs at an incremental cost of USD 9,224 (ICER: USD 15,259 per QALY).

#### Sensitivity analysis results

In one-way sensitivity analyses, the cost-effectiveness of pCGM was primarily driven by reductions in acute event rates rather than by the HbA1c treatment effect, which was modest and non-significant at month 9. When SMBG-associated DKA rates were set equal to those in the CGM arm, pCGM was dominated by SMBG (Table 4). Conversely, substituting for the 6-month trial HbA1c treatment effect rendered pCGM cost-effective in the total population and cost-saving in youth. A sensor price discount of 25% was also sufficient to render pCGM cost-saving in youth. Across all other parameter variations, ICERs for pCGM for youth remained below USD 7,900 per QALY gained (Table 4).

**Table 4:** One-way sensitivity results: Arm 2 (periodic CGM)

|  | Total population |  |  |  |  |  |  | Youth |  |  |  |  |  |  |
| --- | --- | --- | --- | --- | --- | --- | --- | --- | --- | --- | --- | --- | --- | --- |
| Parameter | Total Costs (USD) |  |  | QALYs |  |  | ICER (USD/ QALY) | Total Costs (USD) |  |  | QALYs |  |  | ICER (USD/ QALY) |
|  | CGM | SMBG | Difference | CGM | SMBG | Δ |  | CGM | SMBG | Δ | CGM | SMBG | Δ |  |
| Base case | \$36 765 | \$34 626 | \$2 139 | 7.98 | 7.95 | 0.04 | \$56 128 | \$27 902 | \$27 362 | \$540 | 6.81 | 6.58 | 0.23 | \$2 390 |
| Time horizon* | \$27 194 | \$25 543 | \$1 651 | 6.66 | 6.63 | 0.03 | \$57 222 | \$38 061 | \$36 544 | \$1 517 | 8.18 | 7.81 | 0.37 | \$4 134 |
| ESRD access 0% | \$24 827 | \$23 145 | \$1 682 | 7.98 | 7.95 | 0.04 | \$44 138 | \$20 627 | \$20 772 | -\$145 | 6.81 | 6.58 | 0.23 | -\$644 |
| ESRD access 50% | \$54 671 | \$51 847 | \$2 824 | 7.98 | 7.95 | 0.04 | \$74 113 | \$38 814 | \$37 248 | \$1 567 | 6.81 | 6.58 | 0.23 | \$6 941 |
| DKA cost -15% | \$35 865 | \$33 639 | \$2 226 | 7.98 | 7.95 | 0.04 | \$58 407 | \$26 974 | \$26 213 | \$760 | 6.81 | 6.58 | 0.23 | \$3 367 |
| DKA cost +15% | \$37 665 | \$35 613 | \$2 052 | 7.98 | 7.95 | 0.04 | \$53 849 | \$28 830 | \$28 511 | \$319 | 6.81 | 6.58 | 0.23 | \$1 412 |
| SHE cost -15% | \$36 587 | \$34 402 | \$2 185 | 7.98 | 7.95 | 0.04 | \$57 343 | \$27 713 | \$27 059 | \$653 | 6.81 | 6.58 | 0.23 | \$2 894 |
| SHE cost +15% | \$36 943 | \$34 850 | \$2 092 | 7.98 | 7.95 | 0.04 | \$54 913 | \$28 091 | \$27 665 | \$426 | 6.81 | 6.58 | 0.23 | \$1 886 |
| CGM price -25% | \$36 040 | \$34 626 | \$1 414 | 7.98 | 7.95 | 0.04 | \$37 107 | \$27 315 | \$27 362 | -\$47 | 6.81 | 6.58 | 0.23 | -\$210 |
| CGM price +25% | \$37 490 | \$34 626 | \$2 863 | 7.98 | 7.95 | 0.04 | \$75 149 | \$28 489 | \$27 362 | \$1 126 | 6.81 | 6.58 | 0.23 | \$4 989 |
| Discount rate 0% | \$71 125 | \$67 043 | \$4 082 | 13.34 | 13.24 | 0.09 | \$43 147 | \$40 860 | \$39 718 | \$1 141 | 9.36 | 9.01 | 0.35 | \$3 262 |
| Discount rate 10% | \$23 089 | \$21 712 | \$1 377 | 5.57 | 5.55 | 0.02 | \$62 945 | \$20 136 | \$19 913 | \$224 | 5.22 | 5.06 | 0.15 | \$1 459 |
| SHE rate SMBG -50% | \$36 765 | \$33 927 | \$2 838 | 7.98 | 7.97 | 0.01 | \$231 913 | \$27 902 | \$26 401 | \$1 501 | 6.81 | 6.62 | 0.19 | \$7 891 |
| SHE rate SMBG +50% | \$36 765 | \$35 339 | \$1 426 | 7.98 | 7.92 | 0.06 | \$22 105 | \$27 902 | \$28 227 | -\$325 | 6.81 | 6.55 | 0.26 | -\$1 235 |
| DKA rate = CGM rate | \$36 765 | \$34 317 | \$2 448 | 7.98 | 8.05 | -0.07 | Dominated | \$27 902 | \$25 916 | \$1 986 | 6.81 | 6.85 | - 0.04 | Dominated |
| DKA rate SMBG +50% | \$36 765 | \$35 862 | \$903 | 7.98 | 7.54 | 0.44 | \$2 037 | \$27 902 | \$29 263 | -\$1 361 | 6.81 | 6.31 | 0.50 | -\$2 716 |
| HbA1c Δ month 6 | \$36 343 | \$35 519 | \$824 | 8.11 | 7.64 | 0.47 | \$1 760 | \$27 083 | \$28 841 | -\$1 758 | 6.93 | 6.41 | 0.52 | -\$3 367 |
| *15 years for total population and 100 years for youth |  |  |  |  |  |  |  |  |  |  |  |  |  |  |

One-way sensitivity analyses for Arm 1 (cCGM) demonstrated that results across both populations were most sensitive to DKA rates, HbA1c treatment effect, CGM device price, and discount rate (Supplementary Appendix Figure 1) with all parameter variations either increasing the ICER or not materially altering conclusions.

At a WTP threshold of USD 4,600 per QALY, in the overall population, both pCGM (Arm 2) and cCGM (Arm 1) would require a substantial price reduction (68-73%) to be considered cost-effective, and in the youth subgroup (<20 years), a price reduction of 45% would be necessary to achieve cost-effectiveness for continuous use.

To assess overall parameter uncertainty, we conducted PSA using 100 Monte Carlo simulations. The probability of cost-effectiveness was modest across both arms and subgroups, reflecting substantial uncertainty in the model parameters. For pCGM (Arm 2), the probability of cost-effectiveness at a WTP threshold of USD 4,600 per QALY was 21% in the total population and 53% in youth, rising to 39% and 67% respectively at USD 18,000 (∼3× GDP per capita). For cCGM (Arm 1), the probability was 0-5% at USD 4,600 across both subgroups, increasing to 17% and 51% at USD 18,000 (∼3× GDP per capita). (Table 5, See Supplementary Appendix Figure 2 for Cost-effectiveness planes).

**Table 5:**
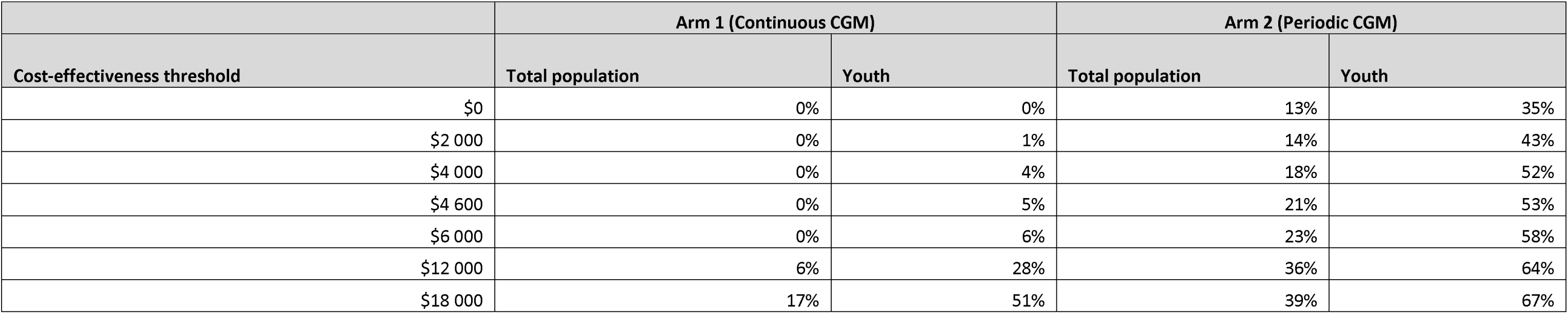
Probability of Cost-Effectiveness by Willingness-to-Pay Threshold, Arm and Population Subgroup (Probabilistic Sensitivity Analysis)

|  | Arm 1 (Continuous CGM) |  | Arm 2 (Periodic CGM) |  |
| --- | --- | --- | --- | --- |
| Cost-effectiveness threshold | Total population | Youth | Total population | Youth |
| \$0 | 0% | 0% | 13% | 35% |
| \$2 000 | 0% | 1% | 14% | 43% |
| \$4 000 | 0% | 4% | 18% | 52% |
| \$4 600 | 0% | 5% | 21% | 53% |
| \$6 000 | 0% | 6% | 23% | 58% |
| \$12 000 | 6% | 28% | 36% | 64% |
| \$18 000 | 17% | 51% | 39% | 67% |

## Discussion

This study evaluated the short- and long-term cost-effectiveness of cCGM and pCGM strategies for people living with poorly controlled T1D in the South African public sector. The within-trial analysis found no statistically significant differences in QALYs or HbA1c across arms. However, when extrapolated over longer time horizons, the model suggested that CGM may provide value for money under certain conditions.

In the within-trial cost-effectiveness analysis, while CGM arms, showed modest numerical gains in utility and glycaemic control, these differences were not statistically discernible. Consequently, no within-trial ICER was calculated. From a cost perspective, cCGM was associated with the highest 9-month direct costs, primarily driven by sensor expenditures, while pCGM offered a more moderate cost profile. As a result, both CGM strategies were dominated by standard of care within the 9-month trial period. These findings underscore the limitations of short time horizons in capturing the full clinical and economic benefits of CGM, especially for chronic conditions like T1D where health and cost impacts accumulate over time.

From a long-term modelling perspective, pCGM was more effective than SMBG; and was associated with only minimal incremental costs among youth. cCGM also yielded meaningful health gains but, due to higher sensor costs incurred higher total costs, resulting in ICERs ranging from USD 15,259 to USD 30,852 per QALY gained. Given the higher costs associated with continuous CGM use, greater reductions in complication incidence are required to generate cost offsets. Although South Africa does not currently use an explicit cost-effectiveness threshold, when these results are interpreted against the Pichon-Rivière et al threshold of approximately USD 4,600 per QALY gained (2025), pCGM is highly cost-effective for the youth subgroup but requires a discount of 68% to be cost-effective for both youth and adults^47^. cCGM may only be considered cost-effective in youth and the total population with a sensor discount of 45% and 73% respectively. Its higher budgetary impact may warrant further prioritisation analysis in resource-constrained settings.

The price reductions required to achieve cost-effectiveness may be more attainable than they appear, particularly given the rapidly evolving CGM market since the trial was conceived in 2022. The ACCEDE trial used the FreeStyle Libre 1 and 2, yet newer market entrants are already available at 20–50% lower cost depending on wear frequency. Public sector volume and price agreements would likely reduce costs further. Notably, our threshold analysis found that a price reduction of just over 25% renders pCGM cost-saving in youth, a reduction already reflected in current market prices before any volume-based negotiation. Taken together, the cost-effectiveness case for CGM in the South African public sector is likely to strengthen considerably as lower-cost devices enter the market, and current model results should be interpreted as conservative.

Our findings sit within the broader evidence on the cost-effectiveness of CGM for individuals with T1D on multiple daily injections, while extending the literature to a LMIC context. A recent systematic review by Jiao et al. reported QALY gains ranging from 0.54 to 3.35 and ICERs between AUD 19,961 and AUD 149,634 across four key studies - Wan et al. (US), Roze et al. (UK and Canada), and Chaugule et al. (US) - all conducted in high-income settings and using higher-cost rtCGM devices such as Dexcom^12–14,18^. For example, the Canadian analysis by Roze et al. estimated annual sensor costs at CAD 3,588, almost double the annualised isCGM device costs in our study, contributing to higher ICERs despite similar health gains. In contrast, the FLASH-UK trial, which evaluated isCGM vs SMBG in adults with T1D, reported an ICER of GBP 4,477 per QALY and found isCGM to be cost-saving in high-risk subgroups. Our pCGM arm, using a lower-cost isCGM device in a resource-constrained LMIC setting, notably yields comparable or more favourable cost-effectiveness estimates than many high-income analyses, and suggests that lower-cost CGM technologies used periodically could offer meaningful value for money in LMIC settings, though further evidence on long-term effectiveness and implementation is needed

One-way sensitivity analyses indicate that pCGM’s economic case rests primarily on reductions in acute hospitalisations rather than HbA1c-mediated complication risk. When CGM-associated DKA rates were set equal to SMBG, pCGM was no longer cost-effective in either population, making the magnitude of acute event reduction the central source of uncertainty. Real-world evidence from large European cohorts demonstrates that isCGM is associated with meaningful reductions in acute diabetes events, including a 50% reduction in DKA hospitalisations in adults with T1D, with consistent reductions in SHE incidence reported across both adult and paediatric populations^49–52^. The relative reductions to SMBG observed in ACCEDE fall within this range, supporting the plausibility of the model inputs, however further evidence from comparable contexts would strengthen confidence in these estimates. Conversely, when the HbA1c treatment effect was more favourable by substituting month 6 trial results where the effect was larger and statistically significant, pCGM became cost-saving in youth and cost-effective in the total population, indicating meaningful upside if glycaemic benefits strengthen with improved engagement and education. cCGM remains sensitive to price and treatment effect assumptions and would require substantially higher WTP thresholds or meaningful price reductions to be considered cost-effective.

To our knowledge, this is the first study to evaluate the cost-effectiveness of CGM in a LMIC setting based on locally generated evidence. Using primary data from the South African ACCEDE trial, this analysis provides context-specific insights into the economic and health value of CGM technologies in a public-sector environment. Importantly, it is also the first to examine pCGM use strategies, which may offer a more affordable and scalable pathway to CGM adoption in resource-constrained settings.

### Limitations

This analysis has several limitations. First, resource use was collected through participant recall over the preceding three months, which may introduce recall bias and lead to underestimation of total resource use; however, these limitations are unlikely to differentially affect results across study arms. Secondly, the DEDUCE model did not fully support TIR as an effectiveness input, despite emerging evidence suggesting that TIR and other CGM-enabled metrics may better predict long-term complications^53,54^. Thirdly, risk predictions within the model were further constrained by the use of older risk equations (Sheffield), given the lack of more contemporary T1D data. Fourth, in the absence of historical complication data from the trial, imputations were made using distributions from Bhana et al. (2023), potentially introducing uncertainty^45^. However, these parameters were all varied in the PSA. Lastly, the DEDUCE model does not support dynamic ageing of cohort-specific parameters: neither age-dependent risk transitions as youth move into adulthood nor indirect costs across an ageing cohort. Productivity and caregiver costs were therefore applied at the event level, and the youth time horizon was fixed at 15 years, with the youngest participants (∼4 years) ageing out by model end. Both workarounds are conservative and likely underestimate lifetime benefit in the youth subgroup and indirect costs.

## Conclusion

Although CGM was not cost-effective in the short-term trial-based analysis, long-term modelling suggests that CGM could provide good value for money in South Africa under certain conditions. pCGM emerged as a cost-effective strategy in the modelling analysis for youth. These findings suggest that a cautious, targeted introduction of pCGMs could be considered within public-sector diabetes programmes in South Africa and similar LMIC settings. To realise this value, broader adoption and equitable access will likely require manufacturer-led price reductions aligned with public-sector affordability constraints, as well as sustained adherence, structured diabetes education, and delivery within a well-functioning diabetes care ecosystem.

## Data Availability

All data produced in the present study are available upon reasonable request to the authors

## Competing interests

Abbott sponsored registration (Prof Karsas and Prof Rheeder) for the SEMDSA (Society for Endocrinology, Metabolism and Diabetes of South Africa) congress in Durban 2025. Prof JA Dave has received honoraria from Abbott for continuing medical education activities on continuous glucose monitoring.

## Funding

This trial is sponsored by FIND, with a grant from The Leona M. and Harry B. Helmsley Charitable Trust. The grant code is HCT-NCDS01. As per the funding contract between The Leona M. and Harry B. Helmsley Charitable Trust and FIND the funders have reviewed the manuscript before submission with a focus on wording relating to the funding source and do have final decision on decision to publish.

## Acknowledgements

We would like to thank the participants for taking part in the study and sharing their experiences. We would like to thank the study teams at Steve Biko Hospital, Groote Schuur Hospital, and Red Cross Children’s Hospital: Tanya Kemp, Johane Freitas, Ian Ross, William Toet, Sophie Davies-Van Es, Maleeka Abrahams-Kahaar, Hanadi Alganeeb, Wayne S. Rajah, Amith Ramcharan, Deepika Goolab, Razana Allie, Cleon Lekaba, Mary-Jane van Zyl, Buyelwa Majikela-Dlangamandla, Neil Meiring, Tyamkazi Nqekeza, Lisa van Wyk, Laura Symmonds, Lindi Gqamana, Santi Horn, Lindsay Rajah. We would also like to thank colleagues (present and past) at FIND for their contributions: Beatrice Vetter, Sonjelle Shilton, Juvenal Nkeramahame, Dorcas Akach, Berra Erkosar, Mikaela Watson, Mae Linh Dupont, Anjana Tomar, Michael Otieno and Vincent Fiechter.

## Notes

### Clinical Trial

NCT05944718

