## Supplementary Appendix for "Cost-Effectiveness of Continuous Glucose Monitoring for people with elevated HbA1c living with Type 1 Diabetes in South Africa: A Within-Trial and Modeled Economic Evaluation"

### Modelling parameters

Supplementary Table 1: Adjusted mean absolute change in HbA1c (%) from baseline and adjusted mean difference from Arm 3(SMBG) by arm [Confidence Interval, p-value]

| **ARM** | **Total population** | | **Youth (< 20 years)** | | **Fully compliant*** | |
| --- | --- | --- | --- | --- | --- | --- |
|  | **Change from baseline**  **[95% CI]** | **Diff [95% CI],**  **p-value** | **Change from baseline**  **[95% CI]** | **Diff [95% CI],**  **p-value** | **Change from baseline**  **[95% CI]** | **Diff [95% CI],**  **p-value** |
| **Endpoint: Month 9** | | | | | | |
| Arm 1 (continuous CGM) | -1.14 [-1.66, -0.62] | -0.21 [-0.93, 0.51], 0.766 | -1.37 [-2.08, -0.66] | -0.12 [-1.09, 0.85], 0.954 | -1.10 [-1.97, -0.23] | -0.66 [-1.71, 0.38], 0.292 |
| Arm 2 (periodic CGM) | -0.95 [-1.46, -0.44] | -0.36 [-1.07, 0.36], 0.470 | -1.03 [-1.73, -0.33] | -0.24 [-1.21, 0.73], 0.831 | -1.32 [-2.01, -0.63] | -0.60 [-1.49, 0.29], 0.255 |
| Arm 3 (SMBG) | -1.08 [-1.61, -0.55] |  | -1.26 [-2.01, -0.52] |  | -1.09 [-1.63, -0.55] |  |
| **Endpoint: Month 6** | | | | | | |
| Arm 1 (continuous CGM) | -0.89 [-1.44, -0.35] | -0.53 [-1.30, 0.23], 0.228 | -1.04 [-1.79, -0.3] | -0.42 [-1.47, 0.62], 0.611 | -0.95 [-1.87, -0.04] | **-1.14 [-2.23, -0.04], 0.040** |
| Arm 2 (periodic CGM) | -1.16 [-1.67, -0.64] | **-1.13 [-1.88, -0.39], 0.001** | -1.45 [-2.18, -0.71] | **-1.28 [-2.33, -0.24], 0.011** | -1.43 [-2.14, -0.71] | **-1.32 [-2.25, -0.39], 0.003** |
| Arm 3 (SMBG) | -0.51 [-1.05, +0.03] |  | -0.64 [-1.41, +0.14] |  | -0.47 [-1.02, +0.08] |  |

Source: Trial data; Note: Figures bold are significant (p<0.05); Estimates derived from linear regression adjusting for baseline HbA1c and covariates. *Greater than 70% active-time.

Supplementary Table 2: Baseline characteristics of patients used in the model by sub-population

| **Parameter** | **Total population** | **Youth** | **Source** |
| --- | --- | --- | --- |
| Age at diagnosis  Mean (SD) | 10.94 (8.49) | 6.98 (3.64) | Trial data |
| Years since diagnosis at model entry  Mean (SD) | 9.86 (7.35) | 6.29 (3.63) | Trial data |
| Gender (% female) | 60.4 | 63.3 | Trial data |
| Baseline HbA1c level (%)  Mean (SD) | 12.2 (1.8) | 12.6 (1.9) | Trial data |
| Systolic blood pressure (SBP) (mmHg)  Mean (SD) | 127.49 (13.49) | 122.95 (13.14) | Adults: Bhana et al. 2023^1^  Youth: Email correspondence Dr Luvina Dookhony |
| Body Mass Index (BMI) (kg/m2)  Mean (SD) | 24.07 (5.39) | 22.52 (5.03) | Adults: Bhana et al. 2023^1^  Youth: Email correspondence Dr Luvina Dookhony |
| High density lipoprotein (HDL) cholesterol (mmol/L)  Mean (SD) | 1.38 (0.50) | 1.42 (0.52) | Adults: Bhana et al. 2023^1^  Youth: Email correspondence Dr Luvina Dookhony |
| Total cholesterol (mmol/L)  Mean (SD) | 4.54 (1.12) | 4.68 (1.11) | Adults: Bhana et al. 2023^1^  Youth: Email correspondence Dr Luvina Dookhony |
| Current smokers | 14% | 6% | GATS 2021^3^ and StatsSA Mid-year Population Estimates 2025 (assumed smoking prevalence is 0% in 0-14 year olds)^4^. |

Supplementary Table 3: Baseline diabetes-related complications by sub-population

| **Parameter** | **Total population** | **Youth** | **Source** |
| --- | --- | --- | --- |
| Neuropathy | 13.1% | 0.7% | Trial data |
| Peripheral Artery Disease with amputation | 0.4% | 0.0% | Trial data |
| Micro-albuminuria | 14.9% | 8.2% | Trial data: Included micro-albuminuria and 50% general category nephropathy allocated to micro- and macro-albuminuria |
| Macro-albuminuria | 9.6% | 3.4% | Trial data: 50% general category nephropathy allocated to micro- and macro-albuminuria |
| End-stage renal disease (ESRD) | 0% | 0.0% | Trial data |
| Background retinopathy | 0.2% | 0.0% | Trial data: Reworked from breakdown in Bhana et al. 2023^1^ |
| Peripheral retinopathy | 0.1% | 0.0% | Trial data: Reworked from breakdown in Bhana et al. 2023^1^ |
| Macular oedema | 0.0% | 0.0% | Trial data: Reworked from breakdown in Bhana et al. 2023^1^ |
| Blindness | 0.04% | 0.0% | Trial data: Reworked from breakdown in Bhana et al. 2023^1^ |
| Myocardial infarction | 0.0% | 0.0% | Trial data: Assume all cardiovascular complications are heart failure in line with Bhana et al. 2023^1^ (excluding stroke, which was available from the trial data) |
| Stroke | 0.8% | 0.0% | Trial data |
| Angina | 0.0% | 0.0% | Trial data: Assume all cardiovascular complications are heart failure in line with Bhana et al. 2023^1^ (excluding stroke, which was available from the trial data) |
| Heart failure | 2.9% | 2.0% | Trial data: Assume all cardiovascular complications are heart failure in line with Bhana et al. 2023^1^ (excluding stroke, which was available from the trial data) |

Supplementary Table 4: Probability of acute diabetic events by Arm and sub-population

| **Parameter** | **Total population** | **Youth** | **Source** |
| --- | --- | --- | --- |
| Diabetic keto-acidosis (annual probability) | CGM: Arm 1: 20.6%;  Arm 2: 28.1%  SMBG: 31.0% | CGM: Arm 1: 25.0%;  Arm 2: 35.9%  SMBG: 46.3% | Trial data |
| Severe hypoglycemia (annual probability) | CGM: Arm 1:3.2%;  Arm 2: 7.5%  SMBG: 9.6% | CGM: Arm 1: 5.1%;  Arm 2: 9.9%  SMBG: 16.3% | Trial data |
| Diabetic keto-acidosis (mortality probability per event) | 5.71% | 5.71% | Ndebele et al. 2018^5^ |
| Severe hypoglycemia (mortality probability per event) | 0.20% | 0.20% | McCoy et al. 2023^6^ |

Supplementary Table 5: Age-specific mortality rate^7^

| **Age** | **Men** | **Women** | **Age** | **Men** | **Women** | **Age** | **Men** | **Women** |
| --- | --- | --- | --- | --- | --- | --- | --- | --- |
| 0 | 0.0254225 | 0.02015108 | 34 | 0.00650873 | 0.00450388 | 68 | 0.04621822 | 0.02789381 |
| 1 | 0.00391951 | 0.00324457 | 35 | 0.0068347 | 0.00473122 | 69 | 0.04852636 | 0.02980968 |
| 2 | 0.00255855 | 0.00201815 | 36 | 0.00717612 | 0.00497501 | 70 | 0.0509072 | 0.03181314 |
| 3 | 0.00169609 | 0.00127818 | 37 | 0.00753121 | 0.00523331 | 71 | 0.05354175 | 0.03400886 |
| 4 | 0.00116123 | 0.00084094 | 38 | 0.00790123 | 0.00550549 | 72 | 0.05661105 | 0.03652738 |
| 5 | 0.00083247 | 0.00058306 | 39 | 0.00829109 | 0.00578838 | 73 | 0.06023433 | 0.03946122 |
| 6 | 0.00063975 | 0.00043853 | 40 | 0.00870871 | 0.00607618 | 74 | 0.06437147 | 0.04281097 |
| 7 | 0.0005297 | 0.00035868 | 41 | 0.00916323 | 0.00636244 | 75 | 0.06887053 | 0.04652083 |
| 8 | 0.00047676 | 0.00032207 | 42 | 0.00966384 | 0.00664002 | 76 | 0.07330817 | 0.05039478 |
| 9 | 0.00046415 | 0.00031555 | 43 | 0.01021678 | 0.0069056 | 77 | 0.07730987 | 0.05424058 |
| 10 | 0.00048395 | 0.00033283 | 44 | 0.01082599 | 0.00715829 | 78 | 0.08063543 | 0.05791411 |
| 11 | 0.00053428 | 0.00037241 | 45 | 0.01148968 | 0.00740328 | 79 | 0.08351556 | 0.06148651 |
| 12 | 0.0006146 | 0.00043354 | 46 | 0.01220016 | 0.00764467 | 80 | 0.08636581 | 0.06513259 |
| 13 | 0.00072687 | 0.00051656 | 47 | 0.0129521 | 0.00789585 | 81 | 0.08977638 | 0.06918191 |
| 14 | 0.00087298 | 0.0006201 | 48 | 0.01374338 | 0.00817008 | 82 | 0.0940835 | 0.07389934 |
| 15 | 0.00105403 | 0.00074146 | 49 | 0.01457599 | 0.00848174 | 83 | 0.09949483 | 0.07947617 |
| 16 | 0.0012691 | 0.00087562 | 50 | 0.01546127 | 0.00884686 | 84 | 0.10601853 | 0.08594971 |
| 17 | 0.00151477 | 0.00101508 | 51 | 0.01640652 | 0.00927506 | 85 | 0.11342828 | 0.09321674 |
| 18 | 0.00178419 | 0.00115173 | 52 | 0.01743363 | 0.00978594 | 86 | 0.12139927 | 0.10109494 |
| 19 | 0.00207001 | 0.00128355 | 53 | 0.01854927 | 0.01038181 | 87 | 0.1294592 | 0.10926241 |
| 20 | 0.00236526 | 0.00141187 | 54 | 0.01975533 | 0.01105935 | 88 | 0.13720956 | 0.11750846 |
| 21 | 0.00266631 | 0.00154651 | 55 | 0.02104183 | 0.01180169 | 89 | 0.14473404 | 0.12614887 |
| 22 | 0.00296971 | 0.00169724 | 56 | 0.02237504 | 0.01257048 | 90 | 0.15221145 | 0.13517688 |
| 23 | 0.003274 | 0.00187412 | 57 | 0.02374551 | 0.01335058 | 91 | 0.16004028 | 0.14490803 |
| 24 | 0.00357797 | 0.00208168 | 58 | 0.02515031 | 0.01413408 | 92 | 0.16920626 | 0.15682846 |
| 25 | 0.0038801 | 0.00232104 | 59 | 0.02660156 | 0.01492647 | 93 | 0.17944804 | 0.17014698 |
| 26 | 0.00418046 | 0.00258694 | 60 | 0.02815266 | 0.01577038 | 94 | 0.19016263 | 0.18361342 |
| 27 | 0.00447462 | 0.00286305 | 61 | 0.02984587 | 0.0167056 | 95 | 0.20085735 | 0.19687497 |
| 28 | 0.0047615 | 0.0031353 | 62 | 0.03175083 | 0.01780031 | 96 | 0.21144058 | 0.2102724 |
| 29 | 0.00504271 | 0.00339396 | 63 | 0.03390761 | 0.0191027 | 97 | 0.22276606 | 0.22512876 |
| 30 | 0.00532115 | 0.00363261 | 64 | 0.03628362 | 0.02061478 | 98 | 0.23466273 | 0.24097803 |
| 31 | 0.00560496 | 0.0038582 | 65 | 0.03881547 | 0.02231673 | 99 | 0.24677728 | 0.25712285 |
| 32 | 0.00589529 | 0.00407381 | 66 | 0.04137485 | 0.02413377 | 100 | 0.31900576 | 0.28730663 |
| 33 | 0.00619573 | 0.00428646 | 67 | 0.04385918 | 0.02600923 |  |  |  |

Supplementary Table 6: Baseline utilities used in model

| **Parameter** | **Baseline utility** |
| --- | --- |
| Other: 0-25 years | 0.931650602 |
| Other: 25-50 years | 0.880972973 |
| Other: 50-75 years | 0.8165 |
| Other: 75+ years | 0.8165 |

Supplementary Table 7: Utility decrements used in model

| **Parameter** | **Decrement in year of occurrence** | **Subsequent year decrement** | **Source** |
| --- | --- | --- | --- |
| Peripheral Vascular Disease (PVD) | 0.061 | 0.061 | *Beaudet et al.2014*^8^ |
| Peripheral Arterial Disease with Amputation | 0.28 | 0.28 | *Bilir et al. 2018* ^9^ |
| Microalbuminuria | 0 | 0 | *Bilir et al. 2018* ^9^ |
| Macroalbuminuria | 0.048 | 0.048 | *Bilir et al. 2018*^9^ |
| End Stage Renal Disease (Kidney Disease) | 0.083 | 0.083 | *Bilir et al. 2018*^9^ |
| Background Retinopathy | 0.04 | 0.04 | *Bilir et al. 2018*^9^ |
| Peripheral Retinopathy | 0.07 | 0.07 | *Bilir et al. 2018*^9^ |
| Macular Oedema | 0.04 | 0.04 | *Bilir et al. 2018*^9^ |
| Blindness | 0.074 | 0.074 | *Beaudet et al. 2014*^8^ |
| Myocardial Infarction | 0.055 | 0.055 | *Bilir et al. 2018*^9^ |
| Stroke | 0.164 | 0.164 | *Bilir et al. 2018*^9^ |
| Angina | 0.09 | 0.09 | *Beaudet et al. 2014*^8^ |
| Heart Failure | 0.108 | 0.108 | *Beaudet et al. 2014*^8^ |
| Severe Hypoglycemia | 0.055 |  | *Bilir et al. 2018*^9^ |
| Diabetic Keto-acidosis | 0 |  | *Assumption* |
| Fear of hypoglycemia | 0 |  | *Assumption* |
| Fingerstick disutility | 0 |  | *Assumption* |

### Costs: Within-trial and model

Supplementary Table 8 and 9 summarise the unit costs, within-trial resource use, and modelling assumptions applied for each cost category. These cover blood glucose monitoring (glucometers, strips, lancets), medicines, diagnostics, other diabetes-related consumables, CGM devices, healthcare provider visit costs, hospitalisations and acute events (severe and non-severe hypoglycaemia, diabetic ketoacidosis), indirect costs (absenteeism, presenteeism, and caregiver time), as well as complication costs for the model. For the within-trial analysis, all resource use data were extracted directly from the case report forms completed during the study. For the modelling analysis, resource use assumptions are detailed in the table and, where not taken from the trial, are based on expert opinion or literature.

Additionally, prior to trial implementation, diabetes nurse educators received approximately four hours of CGM-specific training to enable device rollout and participant education. These training costs were allocated across the CGM study arms (Arm 1 and Arm 2) on a per-patient basis and were included under the healthcare provider cost category.

At each of the study visits, participants were asked how many days of work they had missed in the previous 3 months due to diabetes, as well as the number of days of work that they performed at less than 50% productivity due to diabetes. This was to quantify productivity losses due to both absenteeism and presenteeism. For caregivers of participants < 18, they were asked to indicate the number of hours they typically dedicate to diabetes care on an average day. As the DEDUCE model does not capture age-related transitions in productivity or caregiving, indirect costs were estimated using an event-based approach. Productivity losses were valued at the minimum daily wage and assigned as follows: one workday lost per day of hospitalisation for diabetic ketoacidosis (DKA) and severe hypoglycaemia events (SHE)^11^.

Unit costs were assigned to these resource outputs using prices sourced from public sources: laboratory test costs from the National Health Laboratory Service price list^12^, staffing costs from the Department of Public Service Administration Salary scales^13^, medicine and supply costs from the Master Health Product List^14^ or National tenders^15^, minimum wage data from the Department of Labour^11^ and the cost of procedures and in/out-patient hospital stays from Uniform Patient Fee Schedule^16^. Where necessary, unit costs were sourced from other publicly available resources and the literature – noted in the table below.

Supplementary Table 8: Unit costs and resource use assumptions and sources by cost category for both within-trial and modelling

| **Cost category** | **Unit cost (USD)** | **Within-trial resource use** | **Modelling resource use** | **Source of unit cost** | **Notes on unit cost** |
| --- | --- | --- | --- | --- | --- |
| CGMs  (*Abbott Freestyle Libre*) | $60.50/sensor;  $58.89/reader;  $1.87 (plaster) | Actual sensors and readers used as per trial data. One plaster used per sensor. | Arm 1: 26 (52/2) sensors (every 2 weeks); Arm 2: 4 sensors/year (8 weeks CGM use, remainder SMBG); one reader per participant (2-year life); one plaster per sensor | Trial records | No extra costs for sensors/readers replaced under warranty; readers assumed to have a 24-month working life (cost annualized for a 9-month period in the within-trial analysis) (communication Abbott).  Each sensor requires a plaster. |
| Supplies | Strips: $0.06  Lancets: $0.03  Glucagon kit: $21.94  Glucometer: $10.72  Batteries: $0.46  Syringes: $0.11  Needles: $0.02  Sharps: $4.44 | For a number of observations there were lancets, but no strips and vice-versa. Imputed for strips or lancets using the average ratio of lancets to strips. This equated to a small number (34/627) of observations for strips. 24/627 observations were NA for strips. Imputed using the average frequency that conduct SMBG.  All other supply quantities were retained as recorded, with no further imputation applied. | Adults (SMBG): 3 strips/day; Children (SMBG): 4 strips/day; Arm 1: 1 strip/day; Arm 2: 8 weeks CGM (1 strip/day) then SMBG use for remainder; Lancets = 4-5/week; Glucagon = 1 kit/year; Glucometer replaced every 2 years; Syringes/Needles: 1/day (half the year) | MHPL 1 March 2025^14^;  NDOH tender^15^  Glucometer price inclusive of battery and quality control solution. | Assumed glucometer working life is 2 years (annualized to a 9-month period). |
| Medicines | Varies | Insulin units calculated based on prescribed daily dose multiplied by duration of time in care. | Insulin and other medicines from trial prescriptions; scaled to 12 months for modelling (12/9 factor) | MHPL 2025^14^;  Private pharmacy (www.clicks.co.za) | For medicines not listed in the MHPL, prices were taken from a private pharmacy (n=8) |
| Diagnostics | Varies;  NHLS HbA1c: $7.54  Private HbA1c: $14.43  Point-of-care NHLS: $14.11 | For HbA1c tests, included tests conducted 10 days before enrolment dates, and up to 3 weeks after the final visit (to take into account delays from blood draw to test being conducted). | 3 HbA1c tests per year (at public sector NHLS price); Other lab costs from trial scaled to 12 months for modelling (12/9 factor) | NHLS 2023/2024 fee schedule^12^; trial records;  Radiology (Imaging) used the UPFS^16^ | NHLS 2023/2024 schedule inflated to 2025 using StatsSA consumer price index^17^ (statssa.gov.za).  The NHLS price for HbA1c is a weighted average of *private* NHLS rate (to ensure results returned on time, $14.11 n=157/792) and public NHLS rate ($5.92). In addition, during a hack on the NHLS, the private sector (Ampath) was used for HbA1c tests ($14.43). 1 site conducted tests on a point of care device (Siemens DCA Vantage) at a NHLS price of $14.11. |
| Healthcare Providers and visits | Varies  $0.28 per participant for CGM training | Includes Nurse Educator sessions as part of trial (used time recorded for Nurse Educator sessions); Replaced Nurse Educator visits > 105min with mean visit time (n=3). For other healthcare provider consultations, assumed a 15min consultation. Study visits included: enrollment, 3,6,9 months.  Professional nurse received CGM training (~4 hours) to support device rollout and participant education. The associated cost was allocated across CGM arms on a per-patient basis. | Healthcare provider cost from trial scaled to 12 months for modelling (12/9 factor) | National salary scales^13^ ; UPFS^16^ | Nurse educator sessions evaluated at a Professional Nurse salary; Day hospital or clinic visits costed as per UPFS with clinic visits evaluated as an outpatient visit fee (facility level 1) with a general medical practitioner ($15.83); Study visits evaluated as an outpatient visit fee (facility level 3) with a specialist medical practitioner ($28.22) |
| Hospitalisation | General ward stay ($152.94-$161.94 per day) | All other non-glucose related hospital admission events costed at Length of stay (LoS) x UPFS rate | Not included | UPFS 2025^16^, MHPL 2025^14^, NHLS price list^12^ |  |
| Terminal care costs | $16,122.28 |  |  | Drenth et al. 2017^18^ | Inflated from 2012 to 2025 using medical services inflation from StatsSA^17^ |
| **Glucose-related hospital admission events** | | | | | |
| Hyperglycemia | Varies, depending on LoS | General ward stay for LoS. Medicine: Insulin; electrolyte and fluid replacement. Laboratory: blood glucose (strips+lancet) 6 hourly, urea+ creatinine+sodium+ potassium 1x daily, 1x TSH, lipids, CRP | Not included | UPFS 2025^16^, MHPL 2025^14^, NHLS price list^12^ | Expert opinion informed resource use |
| Diabetes ketoacidosis | Varies, depending on LoS. Average: $1,999.45 | Assume 3 days in High care ward; rest of LoS in General Ward. Medicines: Insulin; sodium chloride+ potassium chloride; Enoxaparin prophylaxis; Laboratory: U&E and blood gases every 2 hours, glucose (strips+lancet) every hour, ketones twice daily (for first day), following days: glucose 6 hourly, 1x Urine analysis, 1x TSH, lipids, 1x CRP | Avg. 6.4 days stay from trial: 3 days high-care, remainder general ward. | UPFS 2025^16^, MHPL 2025^14^, NHLS price list^12^ | Expert opinion informed resource use.  Note: For the model, productivity and caregiver costs were valued at the minimum daily wage assuming one workday lost per day of hospitalisation for DKA |
| Hypoglycaemia (severe) | Varies, depending on LoS. Average: $1,486.25 | General ward stay for LoS . Medicine: electrolyte and fluid replacement; dextrose. Laboratory: blood glucose (strips+lancet) 6 hourly, insulin levels, urea+creatinine+sodium+ potassium 1x daily. 1x TSH, lipids, CRP | Avg. 7.6 days general ward | UPFS 2025^16^, MHPL 2025^8^, NHLS price list^12^ | Expert opinion informed resource use  Note: For the model, productivity and caregiver costs were valued at the minimum daily wage assuming one workday lost per day of hospitalisation. |
| **Indirect costs** |  |  |  |  |  |
| Absenteeism and presenteeism | Minimum wage/day $12.80 | Absenteeism relates to days of work missed due to diabetes, and presenteeism is evaluated at 50% of the number of days where the individual indicated they work at less than 50% productivity. | Included directly in glucose-related events (see DKA and severe hypoglycaemic events) | National minimum wage 2025^11^ |  |
| Caregiver time | Minimum wage/hour $1.60 | Capped care-giver time at 8-hours a day, or 40-hour week (wage-earning potential). | Included directly in glucose-related events (see DKA, severe and non-severe hypoglycaemic events) | National minimum wage 2025^11^ | Only answered by caregivers of children |

Supplementary Table 9: Unit costs, micro costing assumption and sources by complication for model

| **Complication costs** | **Unit cost (USD)** | **Micro costing** | **Source of unit cost** | **Notes on unit cost** |
| --- | --- | --- | --- | --- |
| Neuropathy | Year 1: $28.41  Subsequent years: $28.41 |  | Bhana et al. 2023^1^ | Inflated to 2025 using medical services inflation from StatsSA^17^ |
| Peripheral arterial disease with amputation | Year 1: $12,786.84  Subsequent years: $0 |  | Bhana et al. 2023^1^ | Inflated to 2025 using medical services inflation from StatsSA^17^ |
| Microalbuminuria | Year 1: $72.17  Subsequent years: $0 | 2 x screening tests per year  2 x GP visit  2 x nurse visit  20mg enalapril once daily for 365 days | UPFS 2025^16^, MHPL 2025^14^, NHLS price list 2018^19^ |  |
| Macroalbuminuria | Year 1: $110.20  Subsequent years: $110.20 | Simvastatin 20mg once daily for 365 days (50% of patients)  Atorvastatin 20mg once daily for 365 days (50% of patients)  Enalapril 20mg once daily for 365 days (100% of patients)  1 x LDL  3 x albumin/creatinine  2 x serum creatinine  1 x urine dipstick and microscopy  2 x potassium  4 x outpatient visits  Calcium carbonate 500mg once daily for 14 days in line with IV iron  Erythropoietin 3 x 20 IU/kg per week for 2 weeks, in line with IV iron (7% of patients)  IV iron 1000mg cumulative dose over 14 days (adults); or 0.5mg/kg IV q4weeks for 12 weeks for pediatrics (14.5% of patients) | UPFS 2025^16^, MHPL 2025^14^, NHLS price list 2018^19^ | Inflated to 2025 using medical services inflation from StatsSA^17^ |
| End Stage Renal Disease (ESRD) | Year 1: $45,931.66  Subsequent years: $22,161.38 |  | Bhana et al. 2023^1^ | Inflated to 2025 using medical services inflation from StatsSA.^17^  Use weighted average of peritoneal dialysis, hemodialysis and renal transplant.  To factor in limited access to dialysis and renal transplant in the public healthcare sector, it was assumed that only 20% of patients with ESRD would be able to access treatment (expert opinion). |
| Background Retinopathy | All years: $53.02 | 1 x retinal photo  1 x ophthalmologist visit | UPFS 2025^16^  Hofman et al. 2014^20^ | Inflated to 2025 using medical services inflation from StatsSA^17^ |
| Peripheral Retinopathy | All years: $133.12 |  | Bhana et al. 2023^1^ – laser treatment | Inflated to 2025 using medical services inflation from StatsSA^17^ |
| Macular Oedema | Year 1: $341.35  Subsequent years: $341.35 | 3 x monthly injections of bevacizumab (anti-VEGF). Assume no vial sharing | MHPL 2025^14^ |  |
| Blindness | All years: $0.00 |  | Bhana et al. 2023^1^ | Inflated to 2025 using medical services inflation from StatsSA^17^ |
| Myocardial Infarction | Year 1: $1,013.83  Subsequent years: $268.12 |  | Bhana et al. 2023^1^ | Inflated to 2025 using medical services inflation from StatsSA^17^ |
| Stroke | Year 1: $2,558.75  Subsequent years: $102.40 |  | Bhana et al. 2023^1^ | Inflated to 2025 using medical services inflation from StatsSA^17^ |
| Angina | Year 1: $3,269.59  Subsequent years: $268.12 |  | Bhana et al. 2023^1^ | Inflated to 2025 using medical services inflation from StatsSA^17^ |
| Heart Failure | Year 1: $2,050.00  Subsequent years: $309.09 |  | Bhana et al. 2023^1^ | Inflated to 2025 using medical services inflation from StatsSA^17^ |
| Myocardial Infarction | Year 1: $2,050.00  Subsequent years: $0.00 |  | Bhana et al. 2023^1^ (assume the same as stroke death within 30 days) | Inflated to 2025 using medical services inflation from StatsSA^17^ |
| Stroke | Year 1: $2,050.00  Subsequent years: $0.00 |  | Bhana et al. 2023^1^ | Inflated to 2025 using medical services inflation from StatsSA^17^ |
| Heart Failure | Year 1: $2,050.00  Subsequent years: $0.00 |  | Bhana et al. 2023^1^ (assume the same as stroke; death within 30 days) | Inflated to 2025 using medical services inflation from StatsSA^17^ |

### Sensitivity Analysis

We conducted deterministic one-way sensitivity analyses to examine the impact of parameter uncertainty on the incremental cost-effectiveness results. Structural uncertainty was explored by varying the time horizon (15-year versus lifetime), applying alternative discount rates (0% and 10%), and modifying assumptions regarding end-stage renal disease (ESRD) access (0% and 50%), which influence long-term complication trajectories and costs. Costing assumptions were examined by varying CGM device price (±25%), DKA and SHE treatment costs (±15%). Treatment effect assumptions were assessed by using the HbA1c change observed at month 6 rather than month 9 from the trial. Clinical parameters included variation in DKA (+50% SMBG, or SMBG DKA rate equal to the CGM DKA rate) and SHE event rates (±50% from SMBG-specific rates).

Supplementary Table 10: Within-trial sensitivity results: UK and Slovenia value set

| **Cost Category** | **N** | **ARM 1 - Mean (SD)** | **ARM 1 - Median (IQR)** | **ARM 2 - Mean (SD)** | **ARM 2 - Median (IQR)** | **ARM 3 - Mean (SD)** | **ARM 3 - Median (IQR)** | **Adj Δ (1 vs 3) [95% CI]** | **Adj Δ (2 vs 3) [95% CI]** |
| --- | --- | --- | --- | --- | --- | --- | --- | --- | --- |
| **Outcomes** |  |  |  |  |  |  |  |  |  |
| QALY | 229 | 0.591 (0.133) | 0.632 [0.545, 0.675] | 0.589 (0.133) | 0.620 [0.568, 0.671] | 0.600 (0.106) | 0.620 [0.538, 0.677] | -0.002 (-0.03, 0.026) | 0.005 (-0.023, 0.032) |
| Utility change | 229 | -0.016 (0.211) | 0.000 [-0.125, 0.057] | 0.022 (0.283) | 0.000 [-0.077, 0.152] | -0.038 (0.251) | 0.000 [-0.162, 0.000] | 0 (-0.073, 0.074) | 0.026 (-0.047, 0.099) |

Supplementary Figure 1: Sensitivity analysis results- Tornado diagram Arm 1 vs Arm 3: Incremental Cost-Effective Ratio

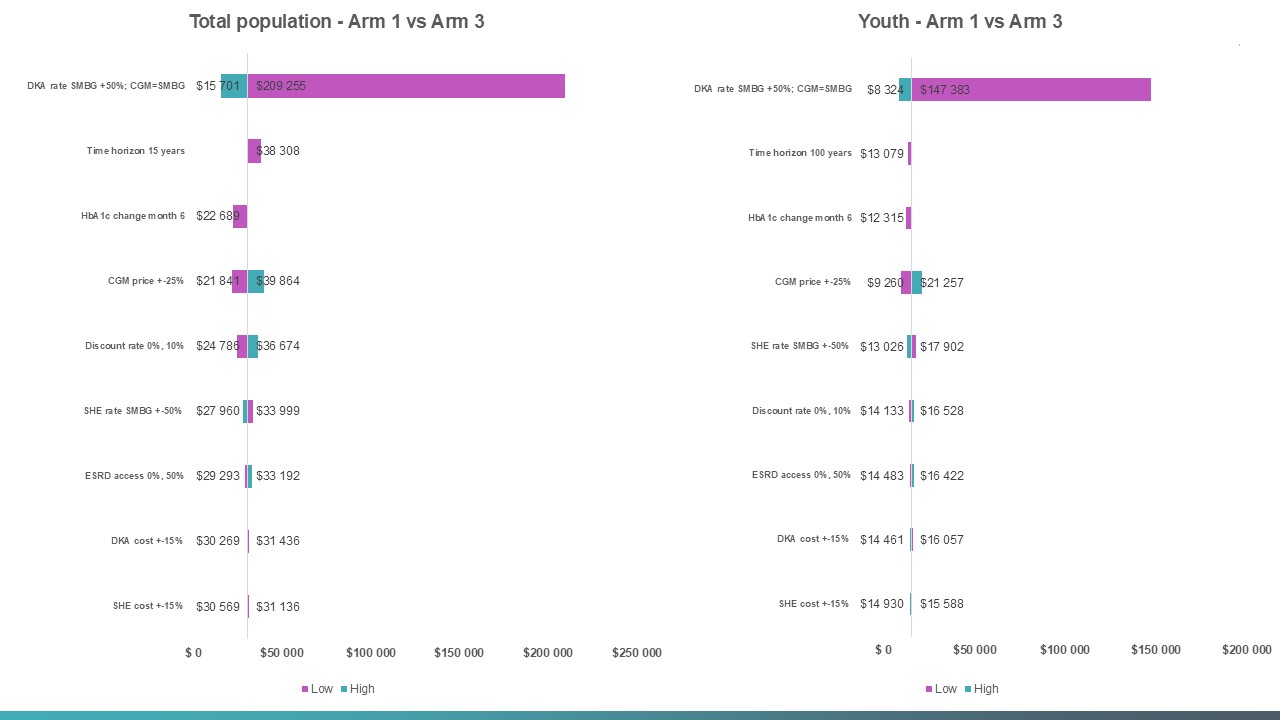

Probabilistic sensitivity analysis (PSA) was undertaken using Monte Carlo simulation, drawing parameters from prespecified distributions reflecting parameter uncertainty: gamma distributions for costs, beta distributions for utilities and probabilities, gamma distributions for utility decrements, Dirichlet distributions for multinomial probabilities, and beta or normal distributions for demographic variables where appropriate.

We initially performed 100 simulations and assessed convergence by visually examining the stability of the mean, lower limit, and upper limit of the net monetary benefit across iterations. If instability in these estimates was observed, additional simulations were undertaken until the estimates stabilised. This approach to assessing convergence is consistent with recommended practice in health economic modelling^21^.

The cost-effectiveness planes show that continuous CGM (Arm 1) was consistently more effective and more costly than standard of care, with simulations concentrated in the north-east quadrant and largely above the USD 4,600 per QALY WTP threshold line (Supplementary Appendix Figure 2). In contrast, periodic CGM (Arm 2) showed a more heterogeneous distribution across the cost-effectiveness plane, with simulations clustered near the origin and spread across multiple quadrants. While a proportion of simulations fell in the south-east quadrant - indicating cost-saving iterations - some simulations also showed negative incremental QALYs, reflecting uncertainty in the direction of the HbA1c effectiveness. Nevertheless, the majority of simulations fell below the USD 4,600 per QALY WTP threshold line, supporting the overall cost-effectiveness of periodic CGM (Supplementary Appendix Figure 2)

Supplementary Figure 2: Cost-effectiveness planes by Arm and sub-population. (WTP = USD 4,600)

Arm 1 - Total population

| Incremental QALYs | Incremental costs (USD) |
| --- | --- |
| 0.485044 | 20390.7 |
| 0.358859 | 13821.3 |
| 0.0113301 | 13601.9 |
| 0.506768 | 17404.8 |
| 0.646009 | 18175.7 |
| 0.555036 | 12499.4 |
| 0.677318 | 16756 |
| 0.317603 | 16427.3 |
| 0.1897 | 29079 |
| 0.518102 | 10021.1 |
| 0.368228 | 16036.2 |
| 0.188357 | 19683.4 |
| 0.247537 | 11618.3 |
| 0.005879 | 15854.4 |
| -0.278559 | 18473.1 |
| 0.137544 | 19053.8 |
| 0.510058 | 15780.2 |
| -0.036145 | 15617.3 |
| 1.11932 | 12409.8 |
| 0.579335 | 21439.3 |
| 1.02321 | 10738.1 |
| 0.579737 | 15726.2 |
| 0.658481 | 15392.6 |
| 0.924447 | 7237.38 |
| 0.338556 | 11887.2 |
| 0.125136 | 14796 |
| 1.45659 | 25921.2 |
| 0.427335 | 13130.7 |
| 0.905442 | 15738.8 |
| 0.674637 | 15679 |
| 0.652954 | 22517.5 |
| 0.54393 | 13622.2 |
| 0.584096 | 15121.1 |
| 0.256027 | 18843.5 |
| -0.0808208 | 10870 |
| 0.544144 | 11210.8 |
| 1.03965 | 13955.8 |
| 0.406271 | 18740.5 |
| 0.871362 | 9544.18 |
| 0.244176 | 13579.7 |
| -0.176379 | 19212.9 |
| 0.724177 | 22976 |
| 0.358715 | 14705.6 |
| 0.555066 | 13892.4 |
| 0.23696 | 17943.8 |
| -0.0595466 | 10779 |
| 0.0554922 | 15001.9 |
| 0.794287 | 14967.3 |
| 0.435691 | 19526.4 |
| 0.393447 | 10258.8 |
| 0.279145 | 15662.1 |
| 0.0387775 | 18830.1 |
| -0.034197 | 20466.3 |
| 0.760438 | 12284.4 |
| 0.563199 | 10973.5 |
| 0.810232 | 18326.6 |
| 0.954585 | 26943.1 |
| 0.177692 | 13967.4 |
| 1.15381 | 18051.5 |
| 0.570425 | 16799.5 |
| 0.743895 | 11079.1 |
| 1.0006 | 12480.6 |
| 0.798229 | 17035.6 |
| 0.345079 | 21198.6 |
| 0.421739 | 10050.2 |
| 0.0788319 | 15638.2 |
| 0.902903 | 18115.3 |
| 0.396761 | 20815.2 |
| 0.11271 | 13249.2 |
| 0.634621 | 11336.6 |
| 0.724936 | 20249.5 |
| 0.402528 | 12122.4 |
| 0.291476 | 13146.3 |
| 0.59717 | 17559.8 |
| 0.238168 | 18518.8 |
| 0.47609 | 11494.5 |
| 0.393847 | 17767.8 |
| -0.0405887 | 15632.8 |
| 0.0136428 | 24385.7 |
| 0.668666 | 11245.7 |
| 0.314901 | 16527.3 |
| 0.560188 | 20222.7 |
| 0.324303 | 7811.46 |
| -0.00771369 | 12145.8 |
| 0.0848292 | 17780.1 |
| 0.292885 | 20173.2 |
| -0.146664 | 15619.6 |
| 0.734626 | 11204.7 |
| 0.833971 | 17034.8 |
| 0.481131 | 18274.1 |
| 0.913706 | 10649.7 |
| 0.418265 | 12761.6 |
| 0.949008 | 17516.3 |
| 0.812136 | 6366.34 |
| 0.570249 | 12144.4 |
| 0.458097 | 10802.3 |
| 0.30447 | 16093 |
| 0.229847 | 11731.2 |
| 0.759261 | 13196.4 |
| 0.37359 | 15988.3 |

| Incremental QALYs | WTP threshold = USD5400 |
| --- | --- |
| -0.5 | -2300 |
| 0 | 0 |
| 2 | 9200 |

Arm 2 - Total population

| Incremental QALYs | Incremental costs (USD) |
| --- | --- |
| 0.0933702 | 4076.31 |
| -0.102821 | 3629.59 |
| -0.425136 | 2612.36 |
| -0.0288732 | 2970.67 |
| 0.38067 | 1095.28 |
| 0.0912389 | 2231.99 |
| 0.237316 | 2554.49 |
| -0.0491425 | 2170.78 |
| -0.296987 | 5746.25 |
| 0.185346 | -568.031 |
| -0.0989966 | 3008.63 |
| -0.345872 | 3593.9 |
| -0.12451 | 2930.74 |
| -0.435137 | 4047.89 |
| -0.687149 | 6224.1 |
| -0.112902 | 2423.74 |
| -0.0327695 | 3313.67 |
| -0.462002 | 3419.3 |
| 0.656954 | -2860.66 |
| 0.269887 | 3589.89 |
| 0.498108 | -279.316 |
| 0.178875 | 1671.78 |
| 0.198942 | 705.665 |
| 0.338297 | -731.47 |
| 0.0474255 | 1376.29 |
| -0.309169 | 2641.93 |
| 0.906288 | 5839.59 |
| 0.13901 | 3185.83 |
| 0.409708 | 2146.96 |
| 0.310225 | 2176.76 |
| 0.182371 | 3531.96 |
| 0.0609443 | 1697.32 |
| 0.12547 | 2324.08 |
| -0.140337 | 3776.1 |
| -0.414699 | 2720.13 |
| -0.0099152 | 608.205 |
| 0.618175 | -1516.12 |
| 0.0884102 | 3021.49 |
| 0.325423 | -1037.1 |
| -0.0282604 | 2104.59 |
| -0.560287 | 4445.35 |
| 0.114826 | 4412.85 |
| -0.0684375 | 3763.74 |
| 0.10813 | 1260.84 |
| -0.180778 | 3978.09 |
| -0.515985 | 4047.6 |
| -0.546343 | 2827.56 |
| 0.384075 | 1553.55 |
| 0.0234988 | 2074.79 |
| 0.0881507 | 626.843 |
| -0.211144 | 3424.8 |
| -0.378423 | 3614.91 |
| -0.483806 | 3115.89 |
| 0.31559 | 2233.43 |
| 0.189908 | -580.658 |
| 0.306119 | 3803.92 |
| 0.169627 | 4499.08 |
| -0.190184 | 3433.28 |
| 0.682105 | 3010.06 |
| 0.264554 | 2553.5 |
| 0.272019 | 1102.53 |
| 0.473188 | -314.026 |
| 0.296423 | 2991.77 |
| -0.112828 | 3712.56 |
| 0.057088 | 483.859 |
| -0.252112 | 2750.23 |
| 0.279148 | 2305.95 |
| -0.00454477 | 3165.01 |
| -0.293027 | 2068.8 |
| 0.351963 | -677.387 |
| 0.263566 | 4136.26 |
| 0.0436842 | 2320.62 |
| -0.0311044 | 1770.42 |
| 0.106087 | 3855.05 |
| -0.0785841 | 2815.52 |
| 0.02146 | 1704.92 |
| -0.232031 | 4687.35 |
| -0.351185 | 2881.17 |
| -0.475973 | 5531.6 |
| 0.379919 | 893.323 |
| -0.103567 | 2321.38 |
| 0.108903 | 3063.43 |
| -0.121832 | -278.446 |
| -0.253255 | 1432.8 |
| -0.269887 | 4667.44 |
| -0.19503 | 4979 |
| -0.58962 | 4590.3 |
| 0.331181 | 376.239 |
| 0.249858 | 1826.77 |
| 0.164459 | 2993.71 |
| 0.50989 | -234.681 |
| 0.221374 | 2155.48 |
| 0.570099 | 2003.3 |
| 0.381923 | -1506.1 |
| 0.197935 | 995.42 |
| 0.146456 | -296.524 |
| 0.0445609 | 2025.44 |
| -0.133693 | 2832.97 |
| 0.232551 | 496.101 |
| 0.0107813 | 3334.13 |

| Incremental QALYs | WTP threshold = USD5400 |
| --- | --- |
| -0.5 | -2300 |
| 0 | 0 |
| 2 | 9200 |

Arm 1 - Youth

| Incremental QALYs | Incremental costs (USD) |
| --- | --- |
| 0.526876 | 13265 |
| 0.49657 | 7977.41 |
| 0.294706 | 8758.32 |
| 0.465526 | 10555.7 |
| 0.730781 | 10818.3 |
| 0.68852 | 6030.53 |
| 0.507886 | 8944.63 |
| 0.438662 | 10262 |
| 0.395236 | 19439 |
| 0.549318 | 4927.33 |
| 0.454295 | 9907.52 |
| 0.390628 | 13189 |
| 0.305455 | 7045.97 |
| 0.197283 | 11266.2 |
| -0.00965272 | 14430.1 |
| 0.285398 | 12770 |
| 0.431282 | 9158.4 |
| 0.160933 | 11737.6 |
| 0.778817 | 3924.41 |
| 0.658768 | 15276.2 |
| 0.95402 | 4075.76 |
| 0.604667 | 8641.45 |
| 0.675803 | 8850.63 |
| 0.878898 | 1184.33 |
| 0.434034 | 7625.65 |
| 0.371884 | 9808.65 |
| 1.14395 | 14497.6 |
| 0.49108 | 8019.4 |
| 0.79765 | 7037.9 |
| 0.821415 | 10210.4 |
| 0.621725 | 13339.7 |
| 0.621087 | 7992.78 |
| 0.679552 | 9276.2 |
| 0.361464 | 12019 |
| 0.148436 | 8198.61 |
| 0.674338 | 6519.64 |
| 0.781323 | 4794.78 |
| 0.610832 | 12493.3 |
| 0.838131 | 2571.66 |
| 0.344526 | 9536.17 |
| 0.201456 | 12637.5 |
| 0.722013 | 13955.5 |
| 0.420805 | 9067.5 |
| 0.593987 | 7881.33 |
| 0.370185 | 13574.4 |
| 0.138112 | 7146.31 |
| 0.228276 | 9064.71 |
| 0.831546 | 9076.72 |
| 0.478086 | 11919.5 |
| 0.503386 | 6575.71 |
| 0.422897 | 10431.1 |
| 0.22591 | 12291.4 |
| 0.106971 | 15790.2 |
| 0.859093 | 6063.23 |
| 0.618338 | 5416.38 |
| 0.846414 | 11556.4 |
| 0.811824 | 16630.6 |
| 0.370547 | 9974.64 |
| 0.956375 | 9233.45 |
| 0.764946 | 11123.1 |
| 0.70259 | 4754.91 |
| 0.8587 | 5202.02 |
| 0.874598 | 11212.7 |
| 0.495052 | 14034.8 |
| 0.568048 | 5480.07 |
| 0.238301 | 10561.9 |
| 0.67539 | 8542.19 |
| 0.583132 | 15095.4 |
| 0.253843 | 7454.39 |
| 0.708246 | 6010.51 |
| 0.784352 | 12645.2 |
| 0.402304 | 5273.53 |
| 0.413485 | 6722.39 |
| 0.684114 | 10667.3 |
| 0.3107 | 12258.8 |
| 0.546547 | 6261.39 |
| 0.499369 | 9657.87 |
| 0.138862 | 10645.1 |
| 0.201231 | 16906.2 |
| 0.752351 | 6065.86 |
| 0.367923 | 10072.4 |
| 0.500993 | 11822.7 |
| 0.447695 | 3006.18 |
| 0.222601 | 8469.12 |
| 0.216383 | 13784.8 |
| 0.395494 | 13354.9 |
| 0.0904068 | 10107.4 |
| 0.726934 | 5632.82 |
| 0.814602 | 9572.56 |
| 0.52821 | 12364.2 |
| 0.830603 | 3275.26 |
| 0.598413 | 9701.12 |
| 0.802081 | 8983.89 |
| 0.787229 | 1951.85 |
| 0.703397 | 6695.52 |
| 0.585909 | 6142.34 |
| 0.436822 | 12117.6 |
| 0.492992 | 7887.93 |
| 0.630789 | 6789.14 |
| 0.526868 | 11353.3 |

| Incremental QALYs | WTP threshold = USD5400 |
| --- | --- |
| -0.5 | -2300 |
| 0 | 0 |
| 2 | 9200 |

Arm 2 - Youth

| Incremental QALYs | Incremental costs (USD) |
| --- | --- |
| 0.203963 | 1950 |
| 0.171573 | 1822.71 |
| -0.0748544 | 1322.69 |
| 0.118205 | 1089.58 |
| 0.421977 | -1599.13 |
| 0.306443 | -198.907 |
| 0.225544 | 152.957 |
| 0.114723 | 397.935 |
| -0.0173648 | 3732.46 |
| 0.232925 | -2212.84 |
| 0.0612588 | 1629.49 |
| -0.0433172 | 2473.64 |
| 0.0128797 | 1601.22 |
| -0.185285 | 3714.61 |
| -0.405047 | 5640.47 |
| 0.0567357 | 666.968 |
| 0.0699963 | 1665.76 |
| -0.225967 | 3590.6 |
| 0.43706 | -5250.52 |
| 0.362489 | 1752.97 |
| 0.54998 | -2428.88 |
| 0.308938 | -763.698 |
| 0.288243 | -1006.91 |
| 0.461479 | -3078.57 |
| 0.132906 | 241.025 |
| -0.0381276 | 1308.67 |
| 0.770445 | 1292.78 |
| 0.206838 | 1084.92 |
| 0.392764 | -882.919 |
| 0.446225 | 308 |
| 0.26606 | 831.714 |
| 0.271802 | 124.316 |
| 0.299102 | 641.863 |
| 0.0488665 | 2107.17 |
| -0.152777 | 3093.76 |
| 0.169178 | -388.381 |
| 0.514328 | -5021.4 |
| 0.294033 | 925.732 |
| 0.455526 | -3278.52 |
| 0.0587648 | 1070.66 |
| -0.150449 | 2746.54 |
| 0.300178 | 1892.66 |
| 0.0538025 | 2372.39 |
| 0.210426 | -505.917 |
| -0.00611574 | 3364 |
| -0.208233 | 3250.89 |
| -0.161309 | 1749.58 |
| 0.436499 | -282.697 |
| 0.129804 | 243.819 |
| 0.187228 | -498.755 |
| -0.0464557 | 2496.43 |
| -0.11529 | 2526.77 |
| -0.233857 | 3754.05 |
| 0.439656 | -155.525 |
| 0.280423 | -2234.22 |
| 0.434148 | 1626.12 |
| 0.354982 | 1964.82 |
| 0.043633 | 2454.95 |
| 0.635052 | -560.104 |
| 0.398908 | 635.602 |
| 0.324856 | -1072.71 |
| 0.476005 | -2476.27 |
| 0.439926 | 1596.12 |
| 0.148869 | 1887.45 |
| 0.235216 | -862.588 |
| 0.00847723 | 1515.64 |
| 0.337102 | -955.203 |
| 0.147887 | 2169.11 |
| -0.0506936 | 555.779 |
| 0.400069 | -2942.81 |
| 0.378641 | 1604.01 |
| 0.174334 | -768.205 |
| 0.142361 | -709.884 |
| 0.294653 | 1595.03 |
| 0.0286456 | 1133.79 |
| 0.171278 | 257.728 |
| 0.0945729 | 2093.19 |
| -0.157661 | 1565.9 |
| -0.163156 | 4093.83 |
| 0.439495 | -1462.19 |
| 0.0510628 | 1021.11 |
| 0.196475 | 557.188 |
| 0.0613406 | -1417.59 |
| -0.0833061 | 540.081 |
| -0.0921593 | 4427.87 |
| -0.0495136 | 3363.29 |
| -0.191462 | 3487.55 |
| 0.39324 | -1425.06 |
| 0.387988 | -31.3852 |
| 0.25165 | 563.287 |
| 0.519922 | -3631.78 |
| 0.358093 | 761.753 |
| 0.474333 | -1089.29 |
| 0.43389 | -3051.81 |
| 0.341884 | -798.97 |
| 0.253167 | -1994.27 |
| 0.20924 | 1114.82 |
| 0.12693 | 1950.86 |
| 0.294865 | -1384.46 |
| 0.159055 | 1806.23 |

| Incremental QALYs | WTP threshold = USD5400 |
| --- | --- |
| -0.5 | -2300 |
| 0 | 0 |
| 2 | 9200 |

### CHEERS checklist

Supplementary Table 11: CHEERS 2022 Checklist^22^

|  | **Item** | **Guidance for Reporting** | **Reported in section** |
| --- | --- | --- | --- |
| **TITLE** | | |  |
| Title | 1 | Identify the study as an economic evaluation and specify the interventions being compared. | Title |
| **ABSTRACT** | | |  |
| Abstract | 2 | Provide a structured summary that highlights context, key methods, results and alternative analyses. | Abstract |
| **INTRODUCTION** | | |  |
| Background and objectives | 3 | Give the context for the study, the study question and its practical relevance for decision making in policy or practice. | Introduction |
| **METHODS** | | |  |
| Health economic  analysis plan | 4 | Indicate whether a health economic analysis plan was developed and where available. | A statistical and health economic analysis plan was developed prior to analysis. The within-trial cost-effectiveness analysis was pre-specified in the SAP. The SAP noted that modelling over a longer time horizon may be undertaken to assess longer-term outcomes; the lifetime modelling analysis reported here was developed subsequently to address this objective. The SAP is available from the authors on reasonable request. |
| Study population | 5 | Describe characteristics of the study population (such as age range, demographics, socioeconomic, or clinical characteristics). | Methods - *Study design*; Results – *Baseline characteristics (*Table 1) |
| Setting and location | 6 | Provide relevant contextual information that may influence findings. | Methods-*Study design*; Published protocol referenced^23^. |
| Comparators | 7 | Describe the interventions or strategies being compared and why chosen. | Methods - *Study design*. Published protocol referenced^23^. |
| Perspective | 8 | State the perspective(s) adopted by the study and why chosen. | Methods - *Within-trial cost-effectiveness analysis*; *Resource use and costs* |
| Time horizon | 9 | State the time horizon for the study and why appropriate. | Methods - *Within-trial cost-effectiveness analysis*; *Model assumptions* |
| Discount rate | 10 | Report the discount rate(s) and reason chosen. | Methods - *Model assumptions*. Alignment with referenced Pharmacoeconomic Guidelines in South Africa. |
| Selection of outcomes | 11 | Describe what outcomes were used as the measure(s) of benefit(s) and harm(s). | Methods – *Measurement of effectiveness*; *Cost-effectiveness analysis*; *Model overview* sections (QALYs and HbA1c); Published protocol referenced^23^. |
| Measurement of outcomes | 12 | Describe how outcomes used to capture benefit(s) and harm(s) were measured. | Methods – *Measurement of effectiveness*; *Cost-effectiveness analysis* (trial HbA1c outcomes and utilities, QALYs) and *Model overview* (utilities and QALY calculation); Results – *Within-trial CEA*. |
| Valuation of outcomes | 13 | Describe the population and methods used to measure and value outcomes. | Methods – *Measurement of Effectiveness; Model overview* (baseline health utilities derived from within-trial EQ-5D analysis; utility decrements from literature; Supplementary Tables S6–S7). |
| Measurement and valuation of resources  and costs | 14 | Describe how costs were valued. | Methods – *Resource use and costs* (micro-costing methods; unit costs from NHLS price list, DPSA salary scales, national tenders, and Uniform Patient Fee Schedule). |
| Currency, price date, and conversion | 15 | Report the dates of the estimated resource quantities and unit costs, plus the currency and year of conversion. | Methods – *Resource use and costs* (all costs reported in 2025 USD after conversion from South African Rand using South African Reserve Bank exchange rates). |
| Rationale and  description of model | 16 | If modelling is used, describe in detail and why used. Report if the model  is publicly available and where it can be accessed. | Methods – *Long-term modelled analysis, Model overview* (DEDUCE diabetes microsimulation model; annual cycles; complication pathways; mortality modelling). Referenced published model^24^ |
| Analytics and assumptions | 17 | Describe any methods for analysing or statistically transforming data, any extrapolation methods, and approaches for validating any model used. | Methods –*Model overview* (model structure, simulation approach, parameter distributions, discounting). |
| Characterizing heterogeneity | 18 | Describe any methods used for estimating how the results of the study vary for sub-groups. | Methods – *Modelled populations* (analyses for total population, youth <20 years). |
| Characterizing  distributional effects | 19 | Describe how impacts are distributed across different individuals  or adjustments made to reflect priority populations. | Methods – *Modelled populations* (stratified analyses by age group to explore differential value). |
| Characterizing uncertainty | 20 | Describe methods to characterize any sources of uncertainty in the analysis. | Methods – *Sensitivity analysis* (deterministic one-way sensitivity analysis and probabilistic sensitivity analysis using Monte Carlo simulation). |
| Approach to engagement with patients and others affected by the study | 21 | Describe any approaches to engage patients or service recipients, the general public, communities, or stakeholders (e.g., clinicians or payers) in the design of the study. | Methods – *Study design* (pragmatic trial embedded in routine public-sector diabetes care; clinical implementation context described). Published protocol referenced^23^. |
| **RESULTS** | | |  |
| Study parameters | 22 | Report all analytic inputs (e.g., values, ranges, references) including uncertainty or distributional assumptions. | Methods – *Resource use and costs*; *Model overview*; Supplementary Appendix Tables S1–S9 (efficacy inputs, utility inputs, parameter values and cost inputs). |
| Summary of main results | 23 | Report the mean values for the main categories of costs and outcomes of interest and summarise them in the most appropriate overall measure. | Results – *Within-trial CEA*; Table 2 (costs and QALYs); Results – *Long-term CEA*; Table 3 (lifetime ICERs and QALY gains) |
| Effect of uncertainty | 24 | Describe how uncertainty about analytic judgments, inputs, or projections  affect findings. Report the effect of choice of discount rate and time horizon, if applicable. | Results – *Sensitivity analysis results*; Table 4 and 5; probabilistic sensitivity analysis (PSA) results and cost-effectiveness planes (Supplementary Appendix Figure 1 and 2). |
| Effect of engagement with patients and others affected by the study | 25 | Report on any difference patient/service recipient, general public, community, or stakeholder involvement made to the approach or findings of the study | Not applicable – no formal patient or public involvement in the design of the economic evaluation beyond participation in the clinical trial. |
| **DISCUSSION** | | |  |
| Study findings, limitations, generalizability, and current knowledge | 26 | Report key findings, limitations, ethical or equity considerations not captured, and how these could impact patients, policy, or practice. | Discussion – *Discussion* and *Limitations* sections. |
| **OTHER RELEVANT INFORMATION** | | | |
| Source of funding | 27 | Describe how the study was funded and any role of the funder in the identification, design, conduct, and reporting of the analysis | Funding statement. |
| Conflicts of interest | 28 | Report authors conflicts of interest according to journal or  International Committee of Medical Journal Editors requirements. | Competing interests’ section. |

12.	National Health Laboratory Service. South Africa. NHLS State Price List 2023.24 [internal document].

19.	National Health Laboratory Service. South Africa. NHLS Price List 2018 [internal document].
